# Challenging the Conventional Treatment Initiation Paradigm: Early Detection of Irreversible Cellular Damage in Cardiac Biopsies of Fabry Disease Before the Formation of Gb3 Inclusion Bodies

**DOI:** 10.1101/2023.08.07.23293800

**Authors:** Chung-Lin Lee, Pei-Sin Chen, Yu-Ying Lu, Ching-Tzu Yen, Chun-Ying Huang, Yen-Fu Cheng, Hsiang-Yu Lin, Chia-Lin Hsu, Yun-Ru Chen, Dau-Ming Niu

## Abstract

**Background:** Fabry disease (FD) is a rare X-linked lysosomal storage disorder that affects multiple organs including the heart. In this study, we investigated if early-stage globotriaosylceramide (Gb3) accumulation, before occurrence of inclusion bodies, could cause significant stress and irreversible damages of the cardiomyocytes in FD patients.

**Methods:** Immunofluorescent (IF) staining was performed on fibroblasts from FD patients with IVS4 mutation and myocardial biopsies from G3Stg/GLAko mice and IVS4 FD patients. The staining targeted interleukin-18 (IL-18), nuclear factor-κB (NF-κB), and inducible nitric oxide synthase (iNOS) as inflammatory and oxidative stress markers. Remarkably, the mouse and human myocardial biopsies selected for this study showed detectable Gb3 accumulation under fluorescence but did not exhibit typical FD (Gb3 inclusion body) pathology. Additionally, alpha-smooth muscle actin (α-SMA) IF staining was conducted to detect fibrosis.

**Results:** The results showed that fibroblasts and cardiomyocytes of mice and patients had significant accumulation of inflammatory markers IL-18 and NF-κB, and oxidative stress marker iNOS. The presence of fibrosis was confirmed in these myocardial biopsies through strong positive staining of α-SMA, despite the absence of typical FD pathology.

**Conclusions:** This study suggests that significant cellular stress and even irreversible damages could exist before the typical pathological changes (inclusion bodies formation) in cardiomyocytes of FD. Based on our findings, it is evident that to prevent irreversible damage and improve the prognosis of patients with FD, treatment should be initiated much earlier than we currently thought.

**Clinical Perspective:** *What Is New?:*

- Evidence suggests significant cellular stress and potential irreversible damage may precede inclusion body formation in Fabry cardiomyocytes.
- Inclusion body formation likely represents a later stage event following intracellular Gb3 accumulation past a critical threshold.

*What Are the Clinical Implications?:*

- These findings indicate a need to re-evaluate conventional Fabry diagnostic criteria.
- There is an emphasis on early therapeutic intervention to mitigate progressive cellular dysfunction, inflammation, and fibrosis.

## Background

Fabry disease (FD) is a rare genetic disorder caused by α-galactosidase A (α-Gal A) activity deficiency, resulting in the accumulation of glycosphingolipids, particularly globotriaosylceramide (Gb3), in various tissues and organs throughout the body, including the kidneys, heart, nervous system, and skin. These sphingolipids accumulations can lead to progressive and multisystemic complications. The symptoms of Fabry disease can vary widely but commonly include chronic pain, fatigue, skin manifestations (such as angiokeratomas), gastrointestinal disturbances, and progressive kidney dysfunction. Left ventricular hypertrophy represents the predominant cardiac manifestation of FD. Additionally, patients may develop arrhythmias, advanced conduction abnormalities, and even heart failure. [1-5].

In Taiwan, the IVS4 + 919G > A (IVS4) variant of GLA is a common mutation associated with late-onset cardiac FD, with a high incidence rate of 1 in 1,600 among the male population [6]. Due to the high prevalence of IVS4 FD in Taiwan, which raised doubts about its pathogenicity, the Taiwan National Health Insurance Administration implemented a treatment guideline requiring IVS4 patients to undergo endomyocardial biopsies to confirm that their hypertrophic cardiomyopathy is indeed caused by FD before they can apply for enzyme replacement therapy (ERT) from the national health insurance. Therefore, our team started to conduct endomyocardial biopsies on these IVS4 patients. Until now, more than 100 patients with IVS4 mutation have received biopsies, and the results have shown typical histopathological changes, such as abundant diffuse cytoplasmic vacuolization (inclusion bodies) on hematoxylin and eosin (H&E) staining and lamellar myelin bodies in electron microscopy, in endomyocardiac biopsies of all ERT naïve IVS4 patients with left ventricular hypertrophy (LVH). This finding indicates that IVS4+919G>A mutation is indeed a real pathogenic mutation [7].

Tissue fibrosis is considered an irreversible event with limited therapeutic intervention available and is a negative prognostic factor for ERT [8]. It has demonstrated that initiating ERT for Fabry cardiomyopathy before the onset of myocardial fibrosis is crucial for achieving sustained improvement in myocardial morphology and function [9]. However, our current study has revealed that in IVS4 patients, even those without LVH, around 38.1% of males and 16.7% of females already had myocardial fibrosis in their hearts, as detected by late gadolinium enhancement heart MRI. In the same study, endomyocardial biopsies were performed on seventeen patients, and all of them had significant classical Fabry pathological changes (abundant inclusions and lamellar myelin bodies) in their endomyocardial biopsies [10].

Unexpectedly, we later discovered that three patients with an IVS4 mutation who exhibited some symptoms/signs of Fabry cardiomyopathy but did not have LVH. Cardiac biopsies were performed, but no typical Fabry pathological changes, such as inclusion bodies or lamellar myelin bodies, were found. However, Gb3 immunostaining revealed substantial Gb3 accumulation, and even extralysosomal Gb3 could be found in one of their biopsies [11]. This finding raises an interesting issue to investigate whether this early-stage Gb3 accumulation (without inclusion bodies or lamellar myelin bodies) could initiate severe inflammatory changes or even irreversible damage to cellular or organ function in these Fabry disease patients.

In this study, our primary objective is to evaluate the severity of inflammatory and oxidative stress in fibroblasts obtained from FD patients with IVS4 mutation compared to non-FD controls. We measured the levels of inflammatory markers such as interleukin-18 (IL-18) and nuclear factor kappa beta (NF-κB) and oxidative stress markers such as inducible nitric oxide synthase (iNOS) to assess the degree of inflammation and oxidative stress and further compare the severity between these two groups. Additionally, we performed myocardial biopsies on crossbred G3Stg/GLAko symptomatic mice and wild-type C57BL/6 mice. We also included the biopsies of the three FD patients mentioned earlier, which exhibited substantial Gb3 accumulation through immunostaining but did not display typical FD pathological changes. Our aim was to compare the severity of inflammatory and oxidative stress between these biopsy groups. Lastly, we performed immunofluorescence staining of alpha-smooth muscle actin (α-SMA) in myocardial biopsies from both mice and patients to detect the presence of fibrosis. The aim of the present study is to determine whether irreversible damage, such as cardiac fibrosis, could exist before the occurrence of typical pathological changes associated with Fabry disease. This information is crucial in selecting an appropriate time for the early initiation of treatment.

## Methods

### Fibroblast culture

Wild-type skin fibroblasts (BCRC number: 08C0011) and skin fibroblasts obtained from IVS4 FD patients (collected from Taipei Veterans General Hospital, IRB number: VGHUST105-G7-6-1) were cultivated in Dulbecco’s modified Eagle’s medium (Thermo Fisher Scientific, Waltham, MA, USA) supplemented with 10% heat-inactivated fetal bovine serum, 1% sodium pyruvate, and 1% L-glutamine in a 100 mm dish. The cells were maintained at a temperature of 37 °C in an atmosphere containing 5% CO2, and the medium was refreshed every 72 hours. For the immunofluorescence assay, the cells were seeded in 6-well plates at a density of 2×10^5^ cells per well and incubated in Dulbecco’s modified Eagle’s medium containing 10% FBS for 24 hours, until they reached 80% confluency.

### G3Stg/GLAko mice preparation

In a previous study, it was observed that G3Stg/GLAko mice exhibited a higher level of Gb3 accumulation compared to GLAko mice. Furthermore, the symptoms displayed by G3Stg/GLAko mice more closely resembled those observed in humans with FD. [12]. Therefore, we chose G3Stg/GLAko mice as our experimental model. Transgenic human Gb3 synthase (TgG3S) mice were prepared as previously described and maintained by breeding with wild-type C57BL/6 mice [13]. The G3Stg/GLAko mouse line was generated by crossbreeding male TgG3S mice with homozygous female GLAko mice [14]. We selected 37-week-old wild-type C57BL/6 and G3Stg/GLAko mice because they correspond to the age range of 30 to 40 years in humans, which is when initial cardiac symptoms typically develop in IVS4 FD patients. Furthermore, the hearts harvested from G3Stg/GLAko mice at this age exhibited Gb3 accumulation through immunostaining but did not display typical pathological changes associated with FD. All procedures followed the principles and guidelines outlined in the Guidelines on Laboratory Animal Use and Management established by the ROC Council of Agriculture. Our studies were approved by the Institutional Animal Care and Use Committee at Taipei Veterans General Hospital.

### Patient Biopsy Samples

Three male patients carrying an IVS4 mutation, displaying symptoms of Fabry cardiomyopathy but without LVH, had their previous cardiac biopsies retrieved for inflammatory and fibrosis immunostaining investigations. None of the patients had received ERT before the biopsy. Despite the absence of typical Fabry pathological features, Gb3 immunostaining revealed significant Gb3 accumulation in the biopsies [11]. The study protocol was approved by the institutional review boards of Taipei Veterans General Hospital (IRB number: VGHUST105-G7-6-1), and all participants provided written informed consent.

### Immunofluorescent staining of fibroblast and myocardial biopsy

Multiple studies have implicated inflammatory markers, including NF-κB and IL-18, in the development of Fabry cardiomyopathy [15-17]. Additionally, increased levels of iNOS have been found to correlate with cardiac dysfunction in FD and are even associated with cardiomyocyte death [18]. Furthermore, α-SMA has been identified as a definitive marker of cardiac fibrosis [19]. In this study, we aimed to investigate whether cellular stress, inflammation, and irreversible damage could already exist in the early stages of Gb3 accumulation in Fabry disease by examining these biomarkers.

For our experimental analysis, we utilized fibroblasts obtained from individuals diagnosed with FD carrying the IVS4 mutation, as well as non-FD controls. Additionally, myocardial biopsies from G3Stg/GLAko mice, wild-type mice, and FD patients with the IVS4 mutation were included. Following the removal of the growth medium, the cells were washed with phosphate-buffered saline (PBS) to eliminate any residual culture media. Subsequently, the cells were fixed using Fixation/Permeabilization solution (BD) for 20 minutes at room temperature. To block non-specific binding sites, the cells were incubated with 5% Fetal Bovine Serum (FBS) for 1 hour at room temperature.

Primary antibodies listed in **Table 1** were applied to the cells overnight at 4°C. After incubation, the cells were washed with 1X Perm/wash solution (BD) to remove any unbound primary antibodies. The cells were then incubated with the corresponding fluorescently labeled secondary antibodies listed in **Table 1** for 2 hours at room temperature. Nuclear staining was performed using 4,6-diamidino-2-phenylindole (DAPI), while lysosomal staining in the Gb3 group utilized lysosomal-associated membrane protein 1 (LAMP-1) as the lysosomal marker. Excess liquid was removed, and the stained cells were mounted on glass slides using Fluorescence-Mounting solution (Dako Omnis, Santa Clara, CA, USA).

**Table 1.** Primary and secondary antibodies.

| Antibodies | Host | Dilution | Structures<br>Identified by<br>Antibodies | Source |
| --- | --- | --- | --- | --- |
| 10745-1-AP<br>(polyclonal antibody) | Rabbit | 1:200 | Nf-kB | Proteintech<br>(Rosemont, IL, USA) |
| 10663-1-AP<br>(polyclonal antibody) | Rabbit | 1:200 | IL-18 | Proteintech<br>(Rosemont, IL, USA) |
| 18985-1-AP<br>( polyclonal antibody) | Rabbit | 1:200 | iNOS | Proteintech<br>(Rosemont, IL, USA) |
| C6198<br>(monoclonal antibody) | Rabbit | 1:400 | α-SMA | Sigma<br>( St. Louis , MO, USA) |
| A2506<br>(Monoclonal antibody) | Mouse | 1:200 | Gb3 | Tokyo Chemical Industry<br>(Tokyo, Japan) |
| Dylight488 Goat<br>GTX213111-04 | Mouse | 1:1000 | Secondary<br>antibody | Genetex<br>(Irvine , CA, USA) |
| Dylight594 Goat<br>GTX213110-05 | Rabbit | 1:1000 | Secondary<br>antibody | Genetex<br>(Irvine , CA, USA) |

Finally, the mounted sections were examined under a fluorescence microscope (Nikon ECLIPSE Ni-E, Tokyo, Japan) equipped with a DP71 digital camera (Olympus) at a magnification of x200 and x600. The acquired images were processed and assembled using Image J software (National Institutes of Health, Bethesda, MD, USA) to facilitate analysis and comparison.

### Quantification of Gb3, NF-kB, IL-18, iNOS and α-SMA accumulations by immunofluorescence signals

To quantify the levels of Gb3, NF-kB, IL-18, and iNOS in IVS4 fibroblasts and cardiomyocytes from patients, as well as in G3Stg/GLAko mice, we employed image analysis techniques. Specifically, we utilized ImageJ software (National Institutes of Health, Bethesda, MD, USA) to calculate the corrected total cell fluorescence (CTCF) of each section. This approach allowed us to determine the fluorescence intensity of the target protein and normalize it to the number of nuclei present in the sample. For each individual sample, a minimum of two sets of image fields containing approximately 100 cells were acquired, averaged, and employed for the computation of CTCF.

The statistical analysis was performed using MedCalc version 22.006 (MedCalc Software Ltd., Ostend, Belgium). The Mann-Whitney U test and Kruskal-Wallis test were employed to evaluate the differences between groups due to the small sample size. Statistical significance was determined with a *p*-value threshold of 0.05, indicating the presence of a significant difference between the compared groups.

## Results

### Gb3, NF-kB, IL-18, and iNOS accumulations in fibroblasts of IVS4 patients

The immunofluorescence results comparing normal human skin fibroblasts with IVS4 FD skin fibroblasts are presented in **Figure 1**. The analysis of Gb3 accumulation revealed a significant increase in Gb3 levels within the fibroblasts of the IVS4 FD group compared to the control group (*p* < 0.001) (**Figure 1a,b**). Notably, similar elevations were observed for NF-kB (**Figure 1c,d**) (*p* < 0.001), IL-18 (**Figure 1e,f**) (*p* < 0.001), and iNOS (**Figure 1g,h**) (*p* < 0.001). These results clearly demonstrate an association between IVS4 FD and significantly elevated levels of Gb3 accumulation, along with enhanced activation of NF-kB, IL-18, and iNOS signaling pathways when compared to normal human skin fibroblasts.

**Figure 1.**
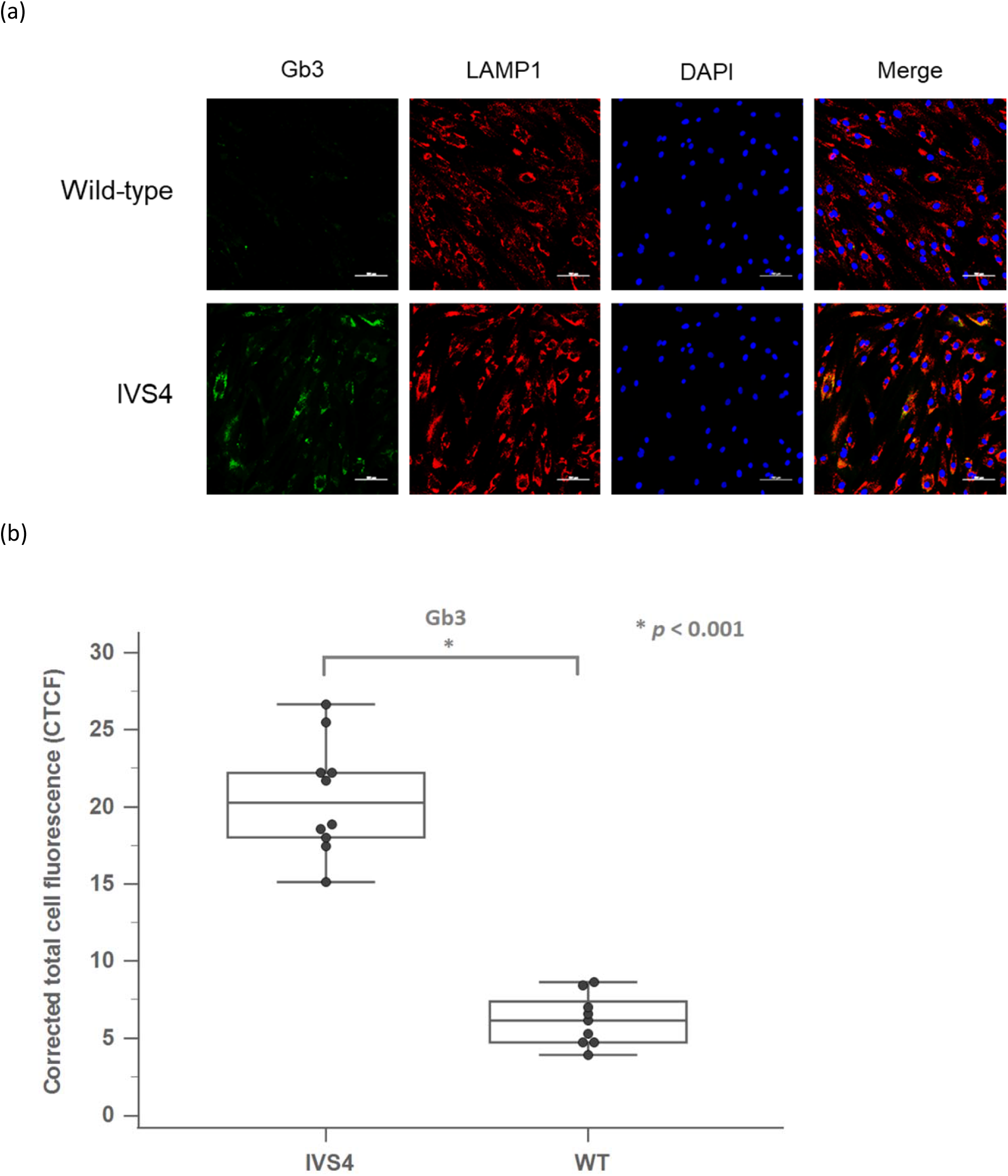
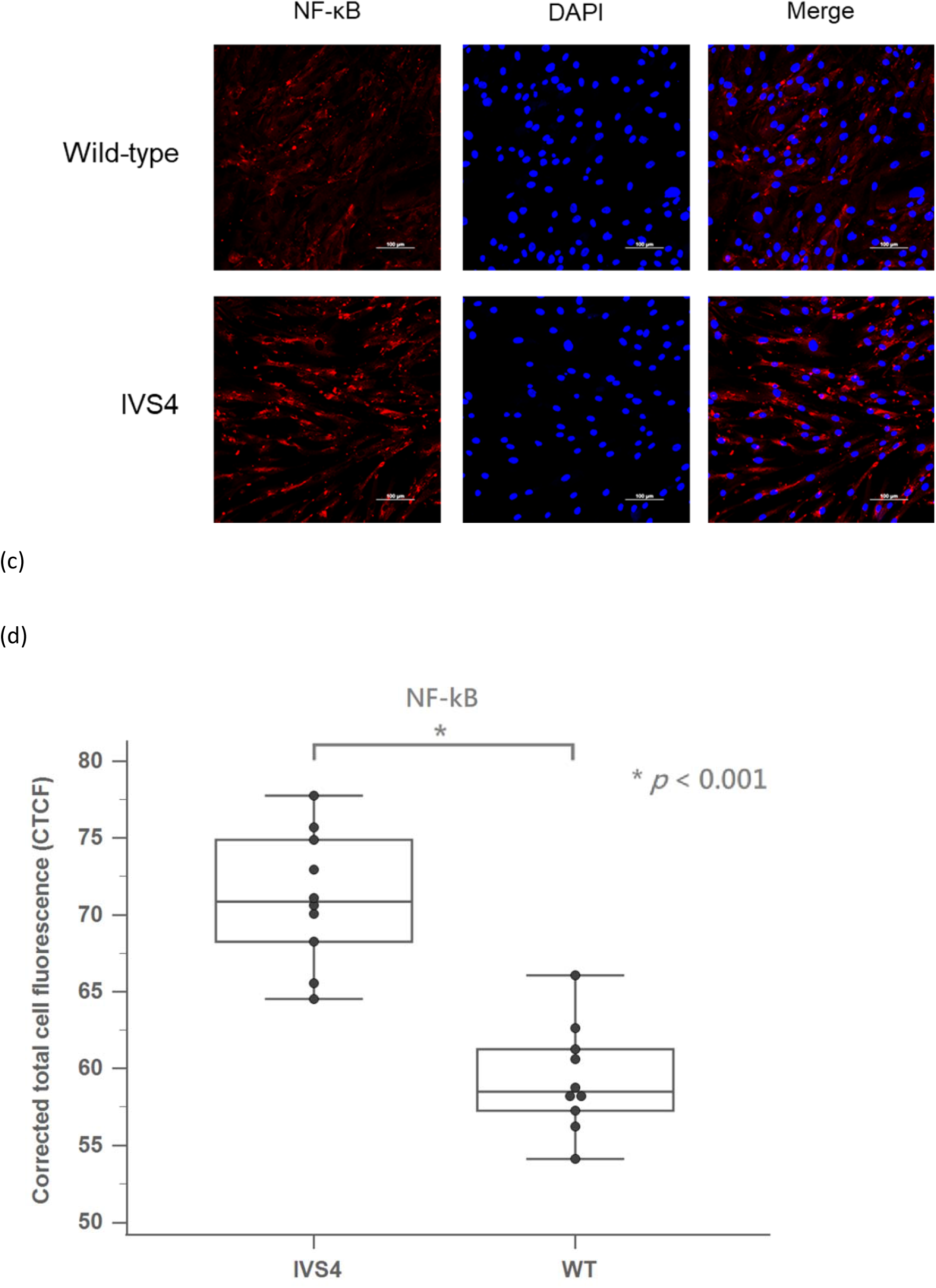
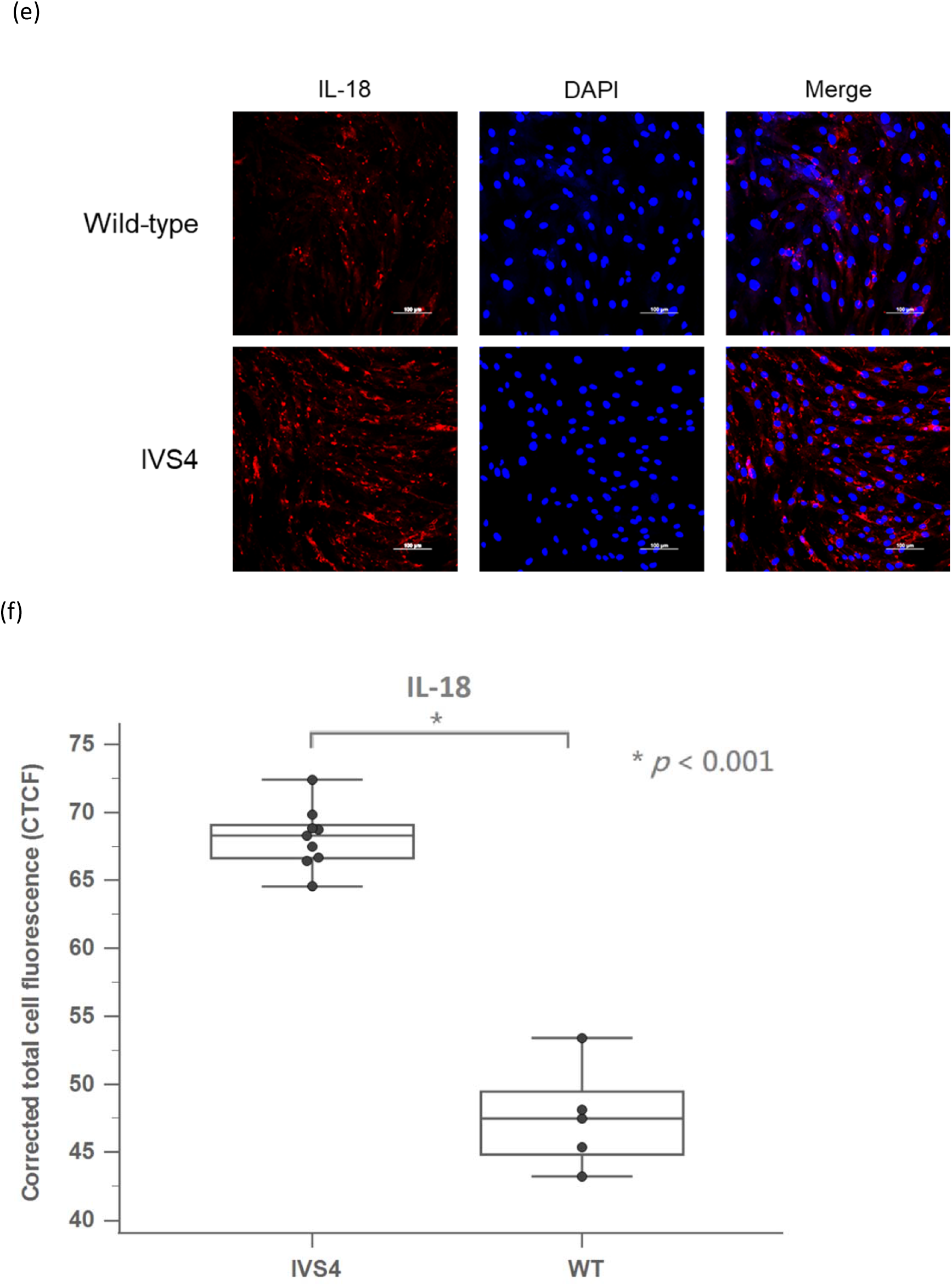
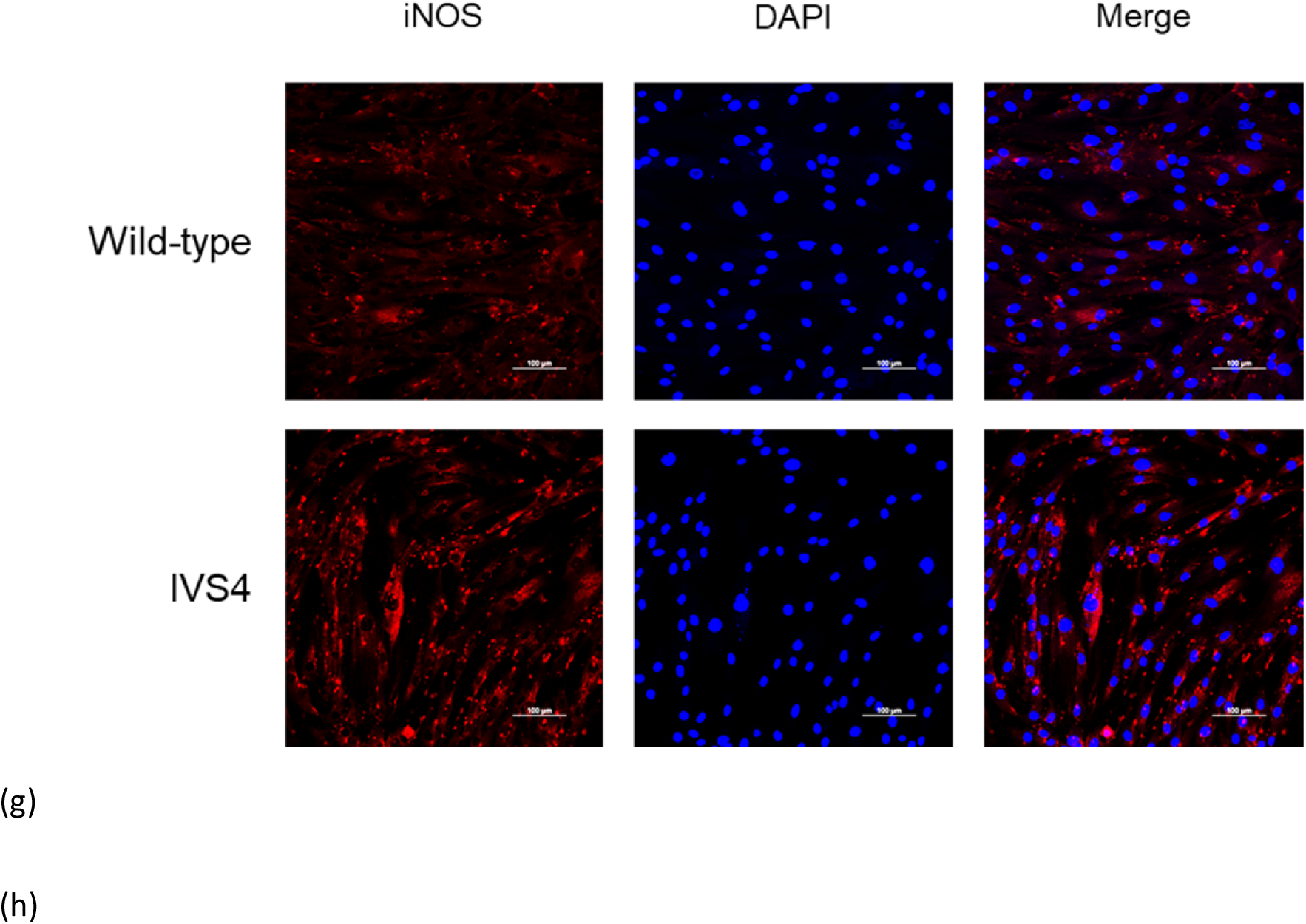
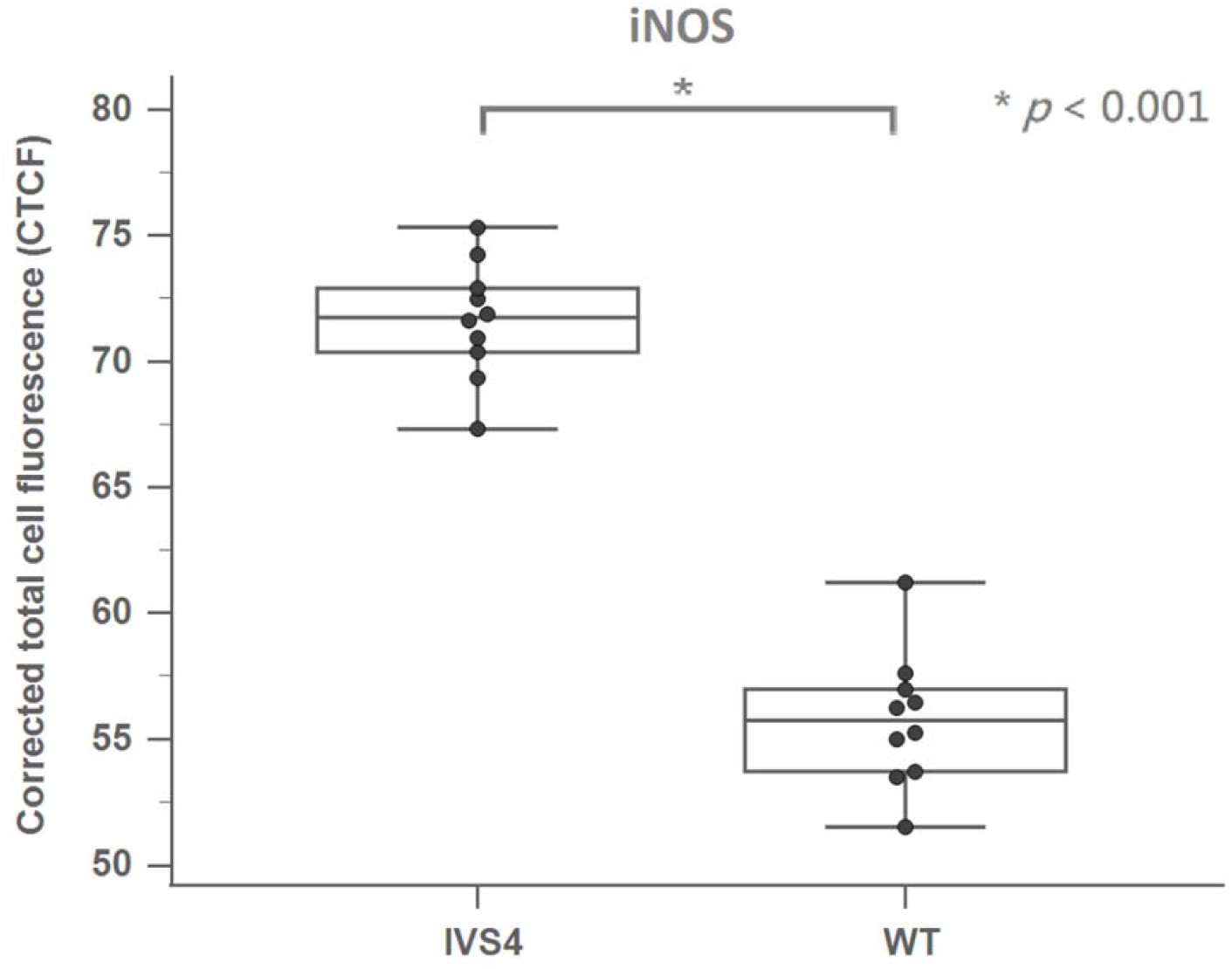
The immunofluorescence result between wild-type and IVS4 FD skin fibroblasts. (a) The immunofluorescence result of Gb3 between wild-type and IVS4 FD skin fibroblasts. (b) The quantification of Gb3 immunofluorescence result between wild-type and IVS4 FD skin fibroblasts. (c) The immunofluorescence result of NF-kB between wild-type and IVS4 FD skin fibroblasts. (d) The quantification of NF-kB immunofluorescence result between wild-type and IVS4 FD skin fibroblasts. (e) The immunofluorescence result of IL-18 between wild-type and IVS4 FD skin fibroblasts. (f) The quantification of IL-18 immunofluorescence result between wild-type and IVS4 FD skin fibroblasts. (g) The immunofluorescence result of IL-18 between wild-type and IVS4 FD skin fibroblasts. (h) The quantification of IL-18 immunofluorescence result between wild-type and IVS4 FD skin fibroblasts.

### Gb3 accumulations in myocardial biopsies of mice and patients with FD

Figure 2a displays the results of Gb3 immunofluorescence analysis conducted on myocardial biopsies from G3Stg/GLAko mice and wild-type mice. Additionally, the Gb3 immunofluorescence analysis findings for myocardial biopsies of three IVS4 patients were previously reported (Figure 3c, f, i) [11]. These images clearly illustrate a significantly higher Gb3 accumulation in cardiomyocytes of both IVS4 patients and G3Stg/GLAko mice compared to wild-type mice. To further quantify the fluorescence intensity, we measured Gb3 in myocardial biopsies from G3Stg/GLAko and wild-type mice. The fluorescence intensity of Gb3 in myocardial biopsies obtained from G3Stg/GLAko mice was significantly higher than that in wild-type mice (Figure 2b) (p < 0.001).

**Figure 2.**
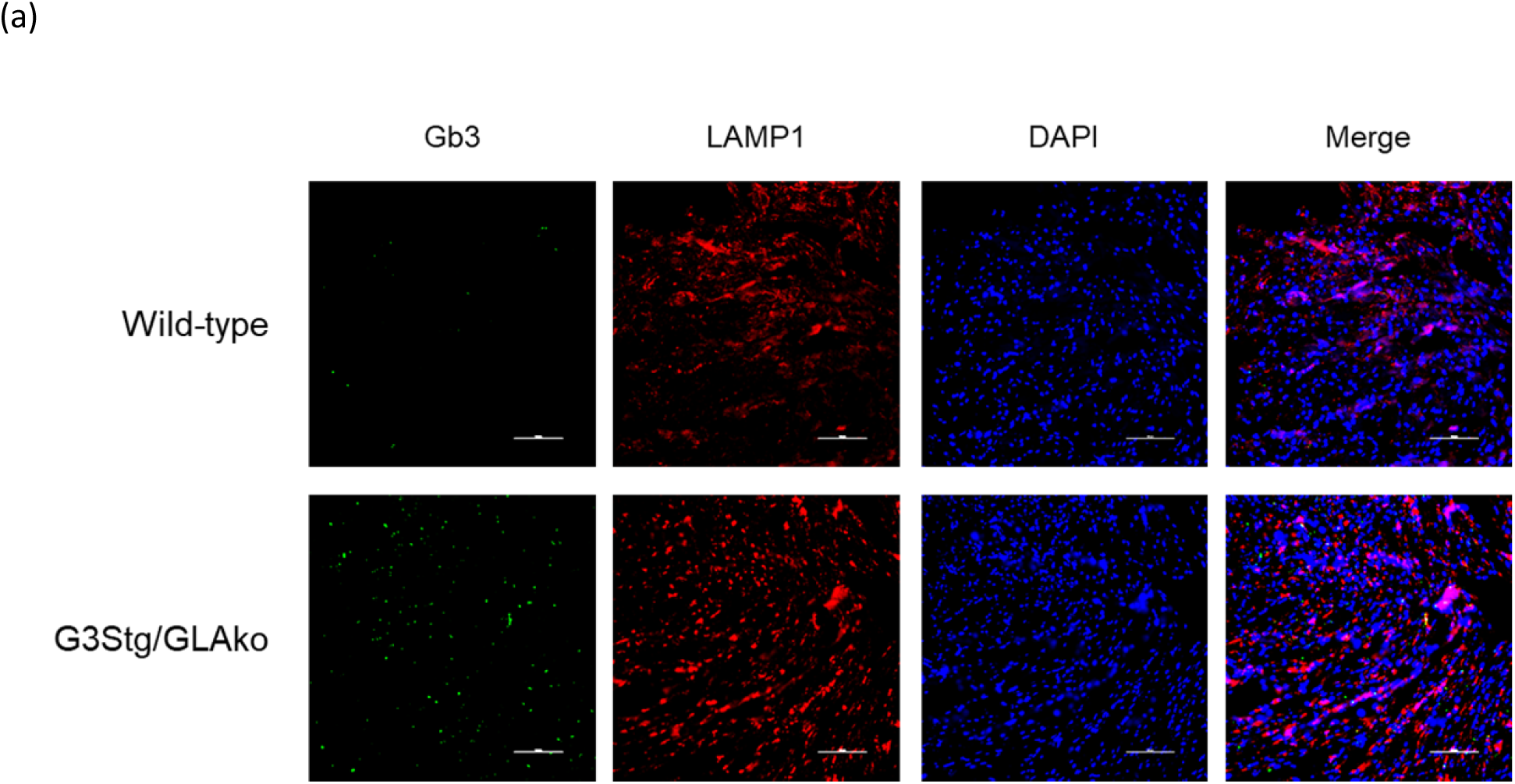
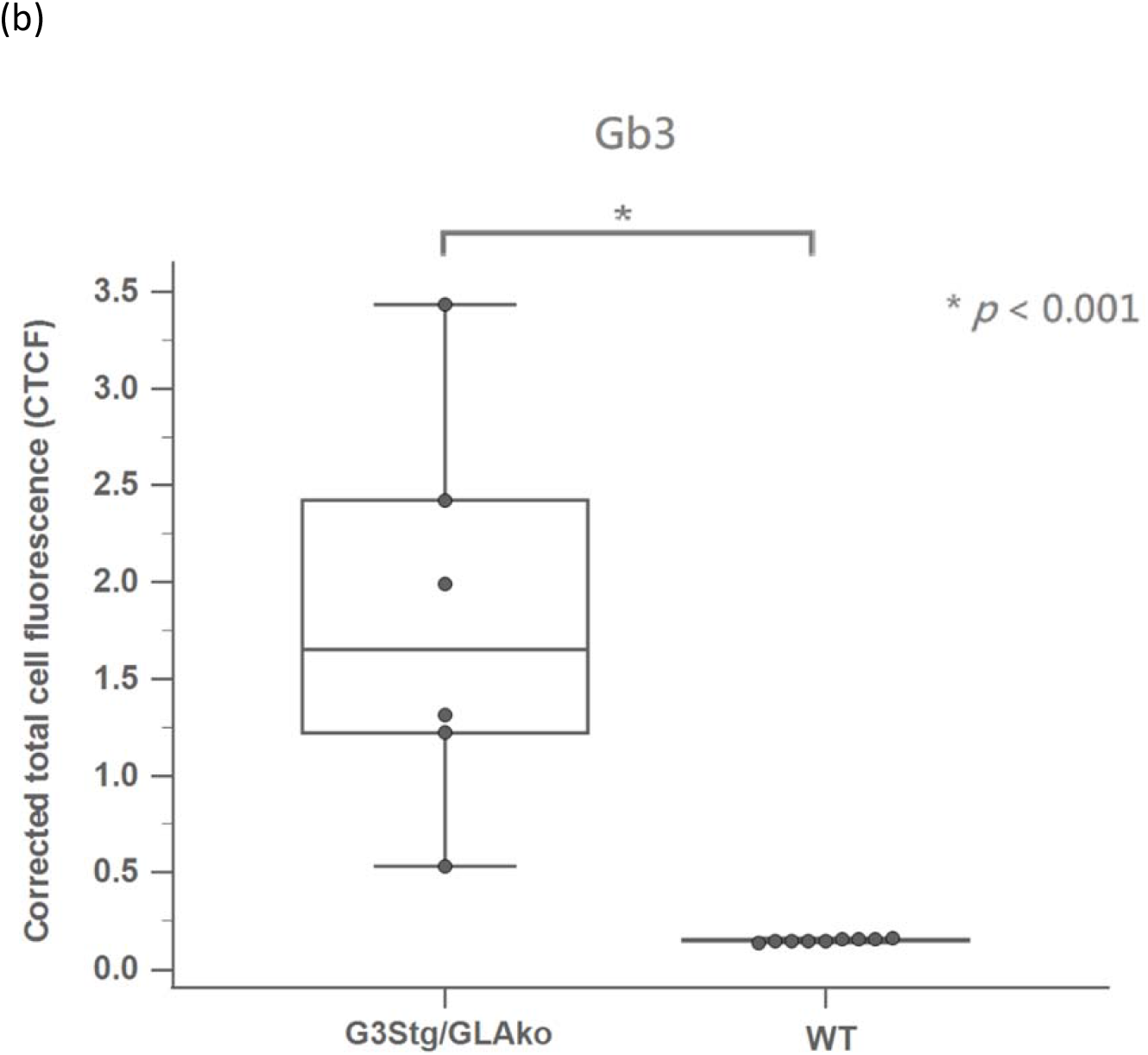
Immunofluorescence analysis comparing myocardial biopsies between G3Stg/GLAko mice and wild-type mice. (a) Immunofluorescence staining of Gb3 in myocardial biopsies from G3Stg/GLAko mice and wild-type mice. (b) Quantification of Gb3 immunofluorescence intensity in myocardial biopsies among G3Stg/GLAko mice and wild-type mice.

### NF-kB, IL-18, and iNOS accumulations in myocardial biopsies of mice and patients with FD

The results of immunofluorescence comparing myocardial biopsies in IVS4 patients with FD, G3Stg/GLAko mice, and wild-type mice are presented in Figure 3. In the NF-kB analysis, NF-kB accumulation was significantly higher in IVS4 patients with FD and G3Stg/GLAko mice compared to wild-type mice (**Figure 3a,b**) (IVS4, *p* < 0.001; G3Stg/GLAko, *p* = 0.005). Similarly, increased IL-18 accumulation was observed in IVS4 patients with FD and G3Stg/GLAko mice compared to wild-type mice (**Figure 3c,d**) (IVS4, *p* < 0.001; G3Stg/GLAko, *p* = 0.009). Furthermore, both IVS4 patients with FD and G3Stg/GLAko mice exhibited higher iNOS accumulation than wild-type mice (**Figure 3e,f**) (IVS4, *p* < 0.001; G3Stg/GLAko, *p* = 0.004). These findings demonstrate significant elevated levels of NF-kB, IL-18, and iNOS accumulations in the early stage (before inclusion bodies formation) of Gb3 accumulation in cardiomyocytes in both IVS4 patients with FD and G3Stg/GLAko mice.

**Figure 3.**
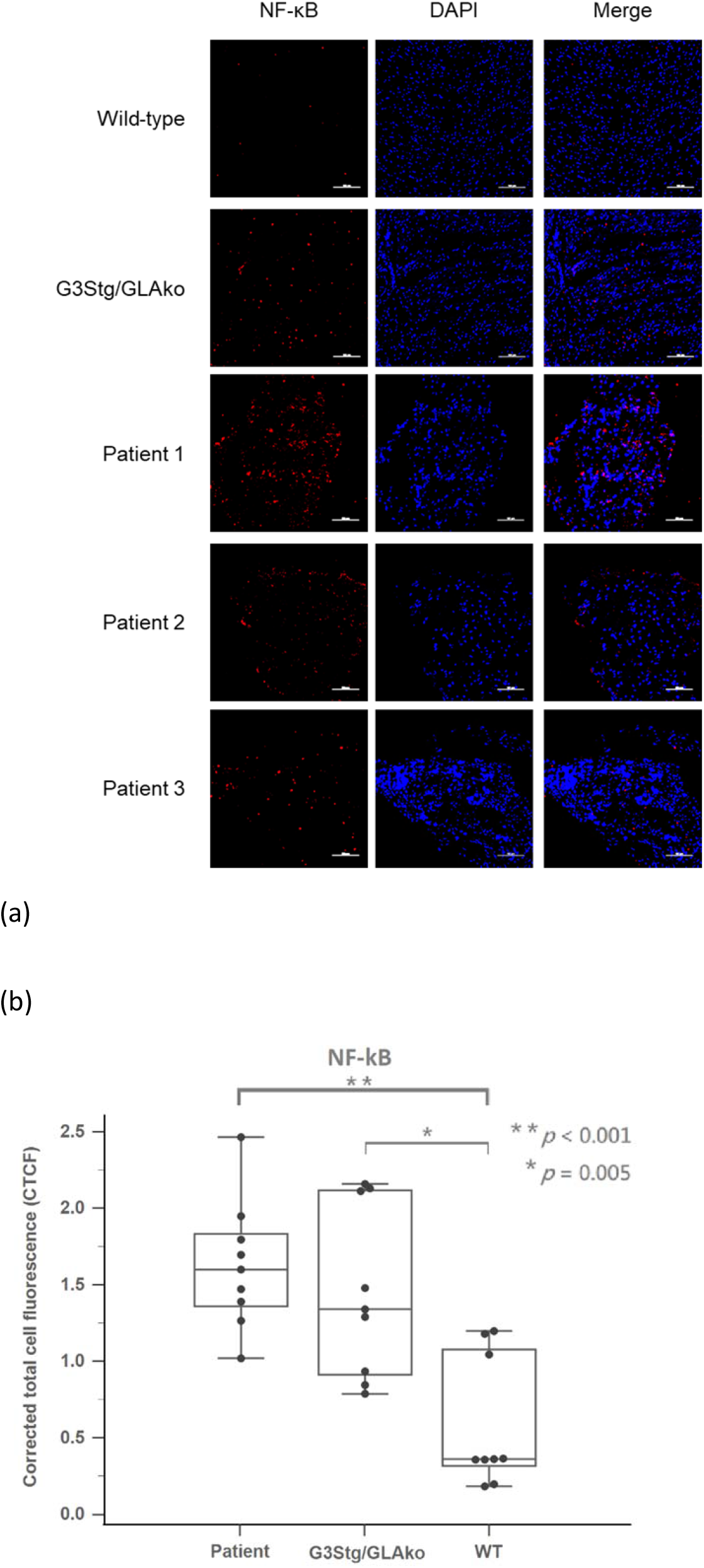
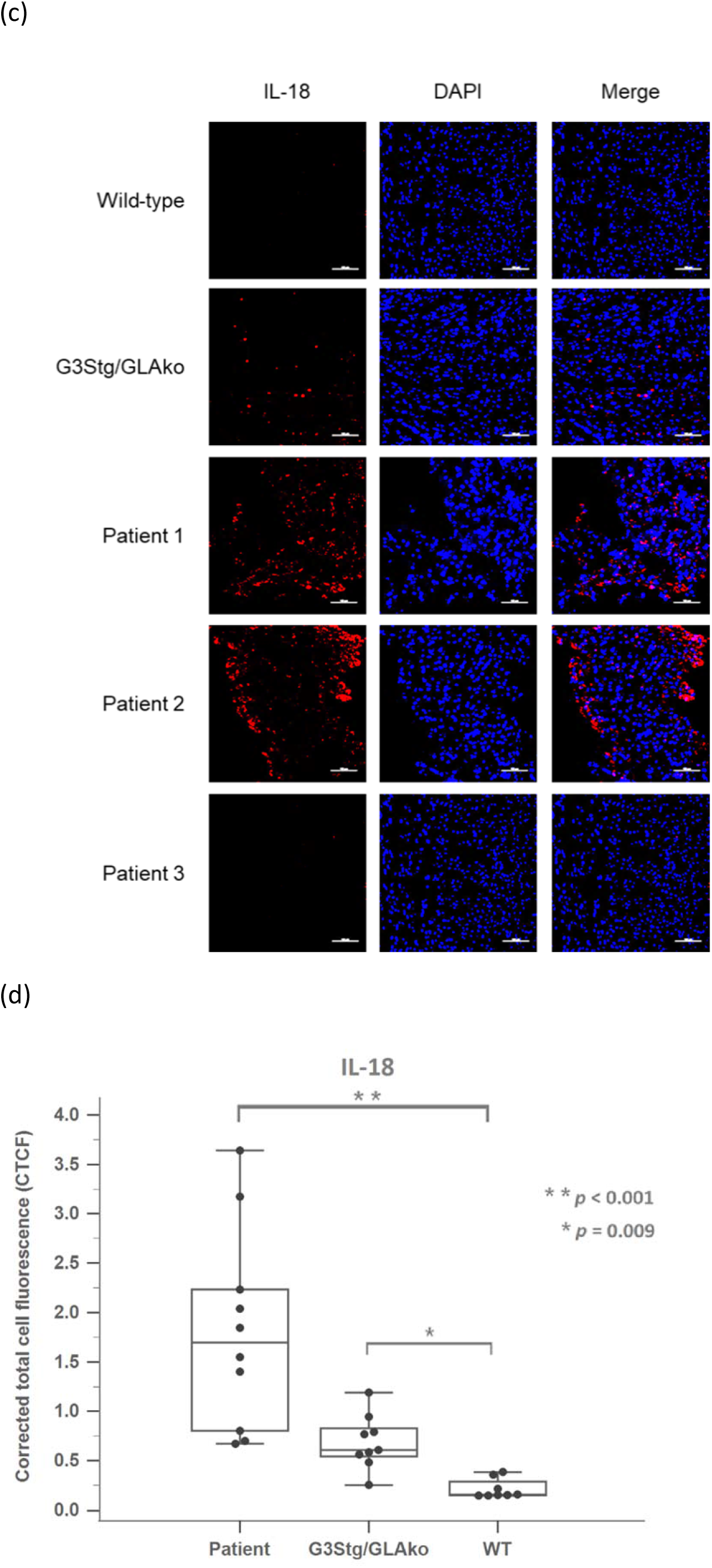
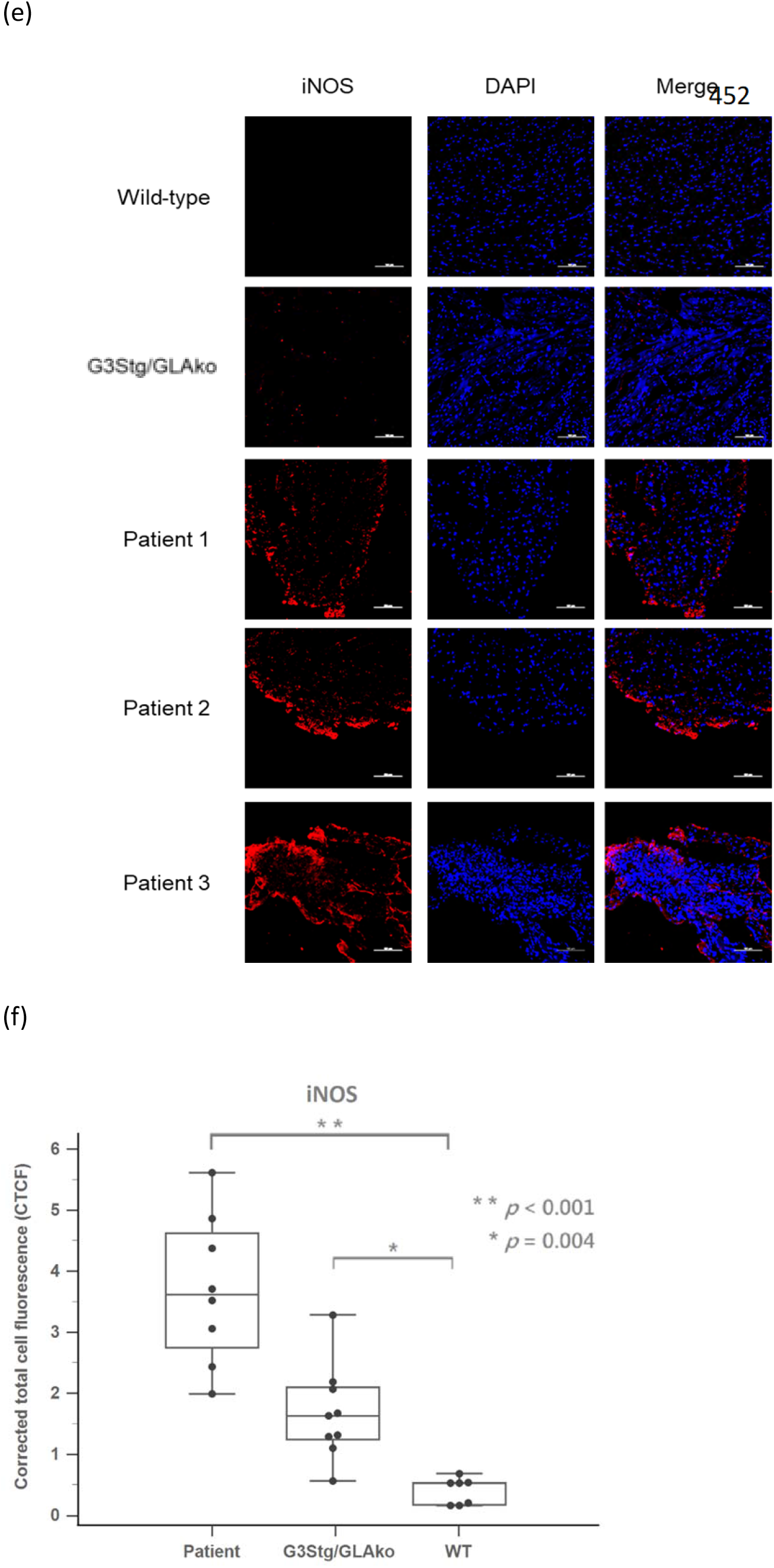
The immunofluorescence result of myocardial biopsies between IVS4 patients with FD, G3Stg/GLAko mice, and wild-type mice. (a) The NF-kB immunofluorescence result of myocardial biopsies between IVS4 patients with FD, G3Stg/GLAko mice, and wild-type mice. (b) The quantification of NF-kB immunofluorescence result between the myocardial biopsies in IVS4 patients with FD, G3Stg/GLAko mice, and wild-type mice. (c) The IL-18 immunofluorescence result of myocardial biopsies between IVS4 patients with FD, G3Stg/GLAko mice, and wild-type mice. (d) The quantification of IL-18 immunofluorescence result between the myocardial biopsies in IVS4 patients with FD, G3Stg/GLAko mice, and wild-type mice. (e) The iNOS immunofluorescence result of myocardial biopsies between IVS4 patients with FD, G3Stg/GLAko mice, and wild-type mice. (f) The quantification of iNOS immunofluorescence result between the myocardial biopsies in IVS4 patients with FD, G3Stg/GLAko mice, and wild-type mice.

### α-SMA accumulations in myocardial biopsies of G3Stg/GLAko mice and IVS4 patients

The results of α-SMA immunofluorescence comparing myocardial biopsies in IVS4 FD patients, G3Stg/GLAko mice, and wild-type mice were presented in Figure 4a. α-SMA expression was significantly higher in IVS4 FD patients and G3Stg/GLAko mice compared to wild-type mice (Figure 4b) (IVS4, *p* < 0.001; G3Stg/GLAko, *p* = 0.032). These findings indicate the presence of fibrosis in the early stage of Gb3 accumulation in cardiomyocytes in both IVS4 FD patients and G3Stg/GLAko mice.

**Figure 4.**
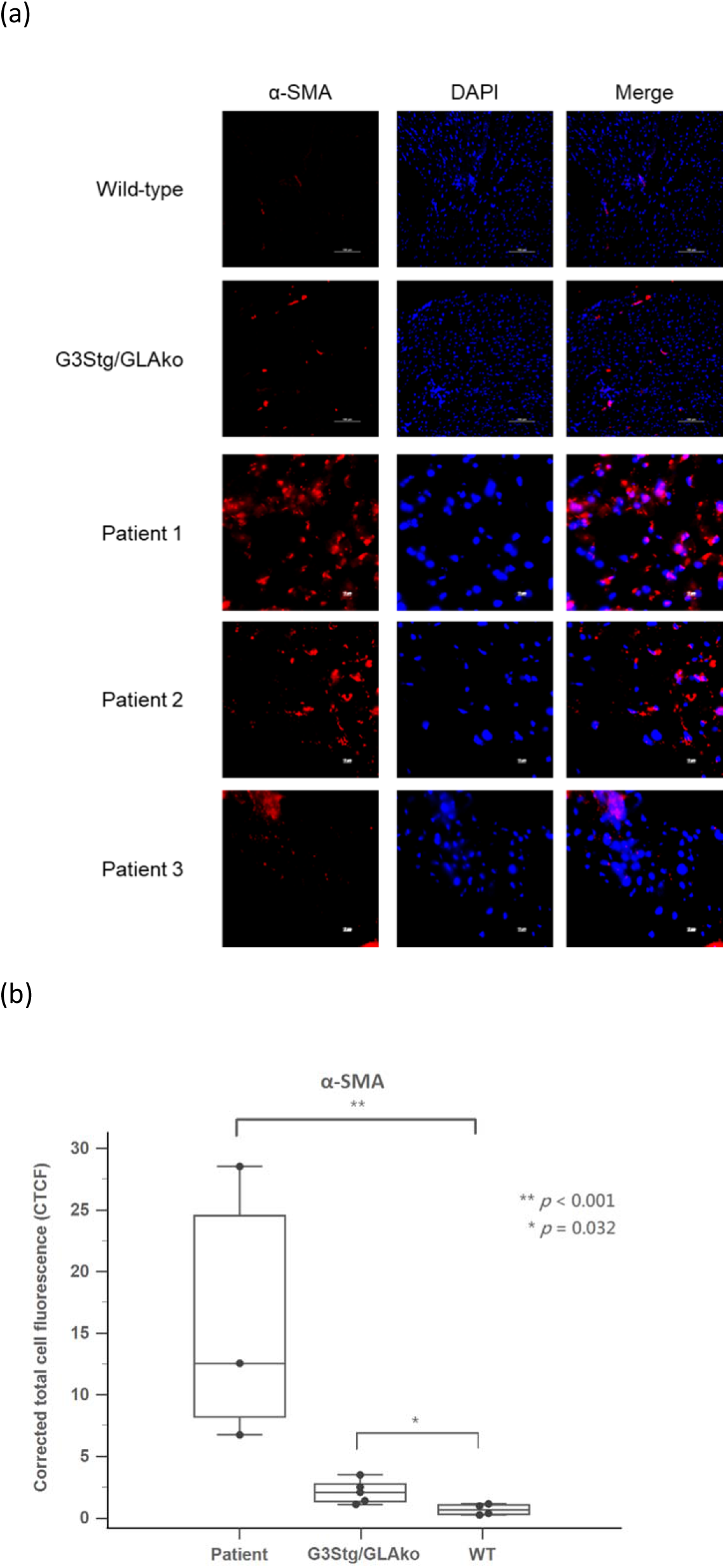
(a) The α-SMA immunofluorescence result of myocardial biopsies between IVS4 patients with FD, G3Stg/GLAko mice, and wild-type mice. (b) The quantification of α-SMA immunofluorescence result between the myocardial biopsies in IVS4 patients with FD, G3Stg/GLAko mice, and wild-type mice.

## Discussion

The findings of this study provide important insights into the early-stage accumulation of Gb3 in FD patients and its impact on cardiomyocytes. Traditionally, the presence of Gb3 inclusion bodies has been considered the hallmark pathological change in FD cardiomyopathy [5]. However, this study challenges this notion by demonstrating that significant cellular stress and irreversible damage may already exist before the typical appearance of inclusion bodies.

The study showed that cardiomyocytes without inclusion bodies from FD patients with the IVS4 mutation exhibited significant accumulation of inflammatory markers and oxidative stress markers. This observation suggests that early-stage Gb3 accumulation can induce significant cellular stress even before the manifestation of detectable pathological changes using current routine examinations, including HE staining and electron microscopic examination. The presence of inflammatory markers (IL-18 and NF-κB) and the oxidative stress marker (iNOS) indicates an activated inflammatory response and increased oxidative stress in FD patients. These findings highlight the potential mechanisms underlying cellular damage in FD and provide future possible targets for therapeutic interventions, such as antioxidant supplementation or immunomodulatory therapy, especially in the early stages of the disease before expensive ERT or chaperone therapy becomes available.

Furthermore, the myocardial biopsies from G3Stg/GLAko mice and three FD patients without typical FD pathological findings demonstrated not only significant cellular stress but also significant fibrosis, despite the absence of inclusion bodies. Fibrosis is a hallmark of cardiac remodeling and can lead to impaired cardiac function [8]. The presence of fibrosis in these samples suggests that irreversible damage can occur in FD patients before the appearance of typical Fabry pathological changes. This finding highlights the importance of early intervention and treatment to prevent irreversible damage and improve patient outcomes.

Regarding the formation of Gb3 inclusion bodies, we believe it to be a result of Gb3 crystallization, which requires a concentration of Gb3 beyond saturation. Therefore, it is plausible that the appearance of inclusion bodies represents a later stage in the accumulation of Gb3. Our study indicates that before the manifestation of inclusion bodies, the intracellular Gb3 level might already be sufficiently high to induce significant inflammatory stress and irreversible tissue damage. These findings have important implications for the understanding of FD progression and the interpretation of histopathological examinations. The routine focus on inclusion bodies (lamellar myelin bodies) alone may underestimate the extent of cellular stress and damage present in FD cardiomyocytes. By solely relying on the presence of inclusion bodies (lamellar myelin bodies) as a diagnostic criterion, the disease may go undetected or be diagnosed at a later stage when irreversible damage has already occurred.

Due to the high prevalence of cardiac-type Fabry disease in Taiwan and our extensive series of myocardial biopsies, we had the opportunity to obtain these three precious biopsy samples. These samples are characterized by exhibiting significant Gb3 accumulation, as observed through immunostaining, yet did not display the typical pathological changes associated with Fabry disease. Interestingly, our findings in these biopsy samples were remarkably similar to those observed in 37-week-old G3Stg/GLAko mice, which corresponds to the human age range of 30 to 40 years. This is the period when patients with IVS4 FD typically develop initial cardiac symptoms. In both the mice and our three IVS4 FD patients who did not exhibit typical pathological changes but showed detectable Gb3 accumulation through immunostaining, we observed significant cellular stress and irreversible damage in the myocardial biopsies. These consistent results not only confirm that early-stage Gb3 accumulation can impair myocardial cells and cause irreversible damage but also highlight the potential of G3Stg/GLAko mice as a valuable research model for studying the impact of early Gb3 accumulation on the heart, especially when this specific type of human myocardial biopsies (without inclusion bodies) is challenging to obtain.

## Conclusions

Our study challenges the conventional understanding of FD progression by highlighting the presence of significant oxidative stress, inflammation, and irreversible fibrotic changes before the appearance of Gb3 inclusion bodies. We propose that inclusion bodies may represent a later stage sign of Gb3 accumulation, occurring after intracellular Gb3 levels have reached a critical threshold. These findings call for a reevaluation of diagnostic criteria and emphasize the importance of early intervention strategies to mitigate cellular stress, inflammation, and fibrosis, ultimately improving the prognosis for patients with FD. Further research is warranted to validate these findings and explore alternative diagnostic markers and therapeutic approaches for the early detection and management of FD.

## Data Availability

All data generated or analyzed during this study are included in this published article.

## Acknowledgments

We acknowledge the participation of study patients and their families.

## Authors’ contributions

D.M.N. was a major contributor in developing the concept. Y.R.C., C.T.Y., and C.Y.H. analyzed the data. C.L.L. performed the experiments assisted by P.S.C., and Y.Y.L.. C.L.L. wrote the manuscript with contributions from H.Y.L., C.L.H. and Y.F.C.. All authors read and approved the submitted version.

## Funding

This study was supported by research grants from the Taipei Veterans General Hospital and University System of Taiwan Joint Research Program (VGHUST105-G7-6-1, VGHUST106-G7-3-1 to C.-L. Hsu and D.-M. Niu) and the Ministry of Science and Technology (MOST), Taiwan (MOST-104-2323-B-010-024 to C.-L. Hsu).

## Availability of data and materials

All data generated or analyzed during this study are included in this published article.

## Ethics approval and consent to participate

The study protocol was conducted according to the Declaration of Helsinki and approved by the institutional reviews boards of Taipei Veterans General Hospital ((VGHUST105-G7-6-1 and VGHUST106-G7-3-1). We obtained written informed consent from all participants.

## Consent for publication

Not applicable.

## Competing interests

The authors declare no conflicts of interest.

## Nonstandard Abbreviations and Acronyms

BD: Fixation/Permeabilization solution and Perm/wash solution
CTCF: Corrected total cell fluorescence
DAPI: 4,6-diamidino-2-phenylindole
ERT: Enzyme replacement therapy
FBS: Fetal bovine serum
FD: Fabry disease
Gb3: Gobotriaosylceramide
H&E: Hematoxylin and eosin staining
IF: Immunofluorescent
IL-18: Interleukin-18
iNOS: Inducible nitric oxide synthase
LAMP-1: Lysosomal-associated membrane protein 1
LVH: Left ventricular hypertrophy
NF-kB: Nuclear factor kappa beta
PBS: Phosphate-buffered saline
TgG3S: Transgenic human Gb3 synthase
WT: Wild type
α-Gal A: α-galactosidase A
GLAko: α-Gal A knockout
α-SMA: Alpha-smooth muscle actin.

## Notes

### Competing Interest Statement

The authors have declared no competing interest.

### Author Declarations

The study protocol was conducted according to the Declaration of Helsinki and approved by the institutional reviews boards of Taipei Veterans General Hospital (VGHUST105-G7-6-1 and VGHUST106-G7-3-1).

